# Effects of Socioeconomic Status in Colorectal Cancer Incidence and Clinical Outcome Differences Among Asian American Populations: A Systematic Review

**DOI:** 10.1101/2025.02.18.25322483

**Authors:** Amy Sakazaki, Austin Lui, Melody Wang, Katherine Ngo, Maria Theresa Lugue, Himani Aligireddy, Maria Nguyen, Shin Murakami

## Abstract

**Introduction:** Colorectal cancer (CRC) is the second leading cause of cancer death in the Asian American and Pacific Islander (API) population, a diverse and rapidly growing minority group in the US. Race, ethnicity, and socioeconomic status (SES) are known to impact outcomes of CRC, but the relationship is unclear in the context of the Asian American cohort and its diverse subgroups. This systemic review aims to gain insight into the relationship between the incidence and clinical outcomes of CRC in the Asian American community.

**Methods:** A systematic literature search was conducted per PRISMA protocol using PubMed, Scopus, EMBASE, Cochrane, CINAHL, and Web of Science databases, accessed on August 13, 2023.

**Results:** Of the 2225 studies identified, a total of 14 studies were included in the analysis. Four studies concluded that there was no association or variable response to SES measurements in CRC incidence in the Asian American population. However, there was evidence that the incidence of CRC varies among Asian American subgroups, using varying measures of SES. In the eight studies that measured mortality or survival as the primary outcome, seven found decreased mortality and increased survival in the API population despite changes in SES. Out of the six studies that measured incidence, four studies found no association with SES. A study found that Chinese Americans experienced a significant decrease in every socioeconomic status (SES) category. Japanese Americans experienced a significant decrease in the lowest SES category, while Koreans and Filipinos experienced a significant increase in both the lowest and highest SES categories. Therefore, grouping various Asian American ethnicities as a single monolithic “Asian” category is misleading.

**Conclusion:** Although the incidence of CRC was thought to be low and decreasing, this review found various trends in different Asian American subset groups. For example, there was a decrease in CRC rates in two ethnic groups and an increase in the other two ethnic groups. The potential causes of these varying CRC incidence rates are likely multifactorial and may include inadequate screening rates, lack of CRC education, and cultural barriers. Further studies are needed to understand these mechanisms. This review recommends a more detailed classification of the API ethnic population but not as a single monolithic entity as Asian. It also emphasizes preventative CRC screening within the API communities due to lower rates of CRC screening among them.

## Introduction

Colorectal cancer (CRC) is the third most common cancer and the third leading cause of cancer deaths in the United States [1]. It is projected that in 2023 an estimated 153,020 individuals will be diagnosed with CRC and approximately 52,550 will die from the disease [1]. Established risk factors include advanced age, genetics, and socioeconomic factors (SES), such as income, education level, insurance, and geographical location [2]. Socioeconomic and cultural factors could also affect potential modifiable risk factors, including smoking, a diet high in processed or red meats and low fruits and vegetables, high alcohol consumption, physical inactivity, and excess body weight [3]. Clinical outcomes of CRC have been improving nationally, likely due to increases in rates of CRC screening [4]. Screening rates also likely represent the ability and accessibility of care.

CRC is more common among the Asian American population in the US and is the second most common cancer [5]. Asian Americans represent 6.2% of the US population and are the fastest-growing racial or ethnic group. According to 2019 data, the Asian American population has increased by 81% in the last two decades [6]. The most prominent subgroups within this population include Chinese, Indian, Filipino, Vietnamese, Korean, and Japanese; however, there are over 20 detailed subgroups listed on the US census [7]. This population embodies a diverse population with an expansive range of languages, histories, cultures, and socioeconomic statuses. Therefore, each group has its own set of health behaviors, genetic and cultural risks, and clinical outcomes. There is a varying degree of acculturation to having a more Western lifestyle and diet, which leads to an increased body weight and decreased physical activity.

Despite the large diversity among the Asian ethnic groups, the medical literature tends to treat them as a single monolithic entity, often together with Pacific Islanders, Asian Americans, and Pacific Islanders (API) [5,8]. This leads to the misleading assumption that all Asian Americans have a comparable health status, despite the inherent diversity among them [9]. For example, it has been reported that Asian Americans, as a monolithic group, have comparable or favorable prognoses compared to other racial or ethnic groups [10-14]. However, when examined as separate ethnic or racial identities, there is considerable variation in incidence trends and outcomes [15]. Mortality rates from CRC also varied, with Native Hawaiians and Southeast Asians having the greatest risk of mortality from CRC; however, Chinese, Japanese, and Indians/Pakistanis had a lower risk [16,17].

The widening of socioeconomic status (SES) disparity in the last several decades has correlated with mortality from all cancers, including CRC, and extends to cardiovascular disease as well [18]. Few studies have examined the effects of SES on clinical outcomes in the context of heterogeneous Asian American ethnicities. With the Asian American population projected to reach 43 million by the year 2050, the factors that affect incidence rates and outcomes of colorectal cancer in the Asian American population must be well characterized to inform preventative measures. Therefore, this systematic review seeks to delineate if SES factors affect clinical outcomes of CRC in the Asian American population. We hope to elucidate which ethnic groups within the Asian American population and what SES factors will impact clinical outcomes of CRC, to focus efforts on cultural and structural implementations for the greatest impact.

## Methods

### Review and Search Strategy

This systematic review was conducted in ordinance with PRISMA criteria and used Covidence software [19,20]. We systematically searched PubMed, Scopus, EMBASE, Cochrane, CINAHL, and Web of Science (last accessed on August 13, 2023). Keywords included Asian Americans, AAPI, Pacific Islander, as well as all Asian subgroups combined with “American” eg., “Vietnamese American”, colorectal cancer, colorectal neoplasm, colorectal tumor, colon cancer, rectal cancer, social determinants of health, healthcare disparities, socioeconomic status, socioeconomic factors neighborhood SES, poverty, social class, income, insurance, occupation, geographic location, literacy, inequality, education, employment, home environment, ethnic enclave, clinical outcomes, treatment outcomes, incidence, mortality, morbidity, vital statistics, progression-free survival, prognosis, intraoperative complications, intraoperative complication, surgical complication, patient outcomes assessment, outcome, critical care outcome, patient care, length of stay, discharge, hospitalization, long term care, palliative care, terminal care, hospice care, rehabilitation, activities of daily living. An example of a complete search strategy can be found in the appendix. Asian American subgroups that were included were Japanese, Chinese, Vietnamese, Asian Indian, Indian, Cambodian, Hmong, Korean, Filipino, South Asian, Indonesian, Pakistani, Malaysian, Bangladeshi, Nepalese, Laotian, Sri Lankan, Bhutanese, Burmese, Taiwanese, Mongolian, Tibetan, Thai, and Singaporean Americans.

### Inclusion Criteria

Criteria for inclusion included the following: peer-reviewed, English language, studies that analyzed incidence rate and/or clinical outcomes (treatment outcomes, incidence, mortality, complications, progression-free survival, etc.) of colorectal cancer patients among Asian American patients and studies that discussed indicators of SES (demographics, socioeconomic status, income, education, geographical location, etc.) as a variable on clinical outcomes. There were no restrictions on the date of publication. Accepted study types included retrospective, prospective, cross-sectional, randomized, nonrandomized, or crossover controlled studies, case series, and case reports.

### Exclusion criteria

Letters, commentaries, conference abstracts, reviews, systematic reviews, meta-analyses; and studies without full text available were excluded.

## Results

We conducted a comprehensive literature search, as described in the Methods section, which identified a total of 2,225 studies. After excluding 80 duplicate entries, we used Covidence software to further screen the titles and abstracts of the remaining studies. Seven independent reviewers assessed the studies to ensure they met the defined inclusion and exclusion criteria. Initially, 38 studies were selected for full-text review by two independent reviewers.

Subsequently, several studies were excluded from consideration: 7 studies did not measure the incidence of colorectal cancer (CRC) or relevant clinical outcomes, 17 studies failed to assess socioeconomic status (SES) indicators, and 17 conference abstracts were disregarded.

Additionally, one study was excluded due to the unavailability of the full text, and two studies were removed because they did not involve populations within the United States. In total, 14 studies were selected for further data retrieval, as shown in Figure 1.

**Figure 1.**
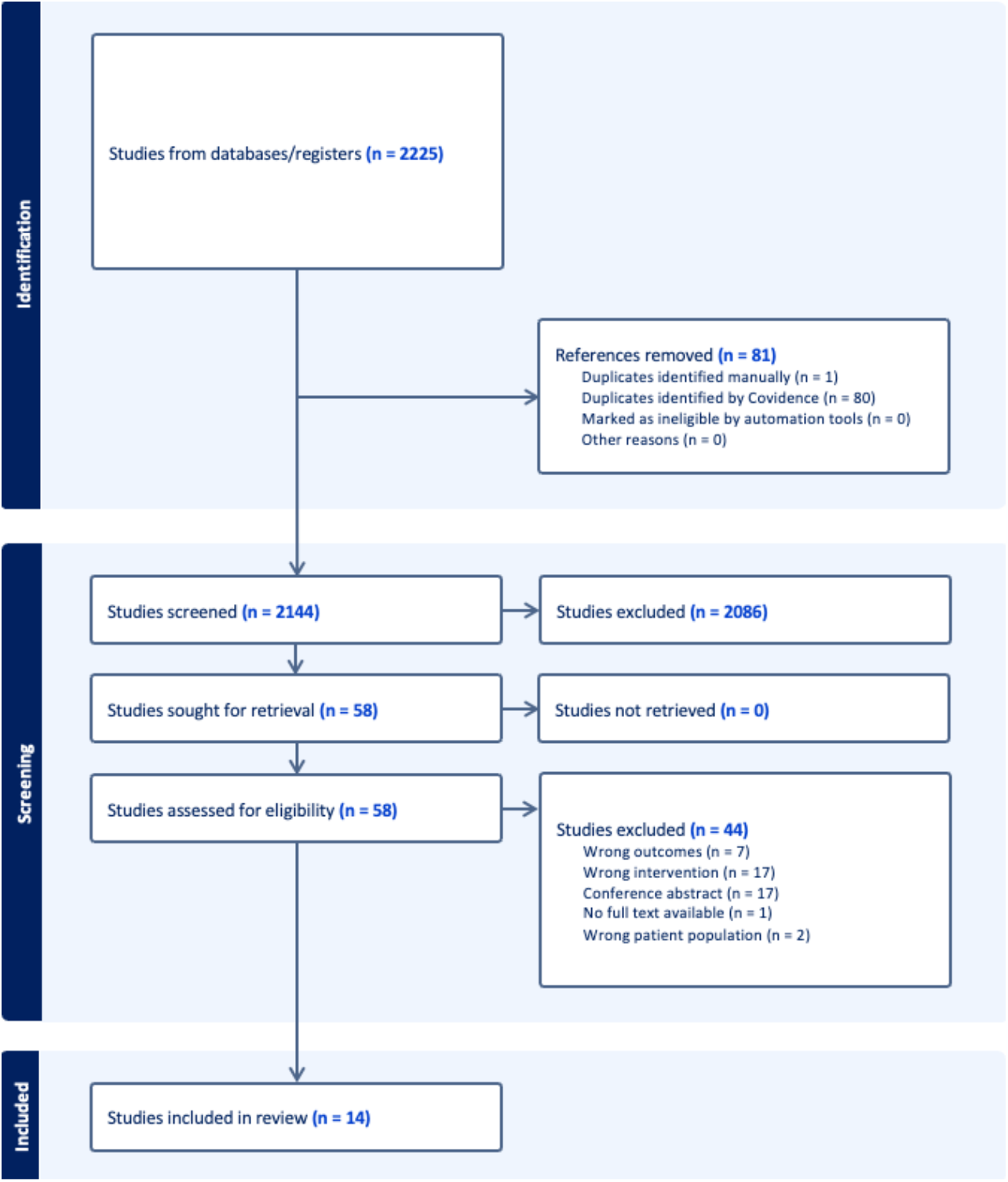
Flow Diagram depicting the study selection process.

The 14 articles identified (Table 1) investigated several socioeconomic variables, including urban vs rural (in two studies), poverty levels (in two studies), SES quintiles or composite SES score (in seven studies), ethnic enclaves (in two studies), and insurance type (in four studies). The outcomes measured in the studies were the incidence rate or rate ratio (in six studies), mortality rate (in five studies), survival rate (in three studies), and laparoscopic surgery (in one study). Additionally four of the 14 articles analyzed at least one specific Asian American subgroup.

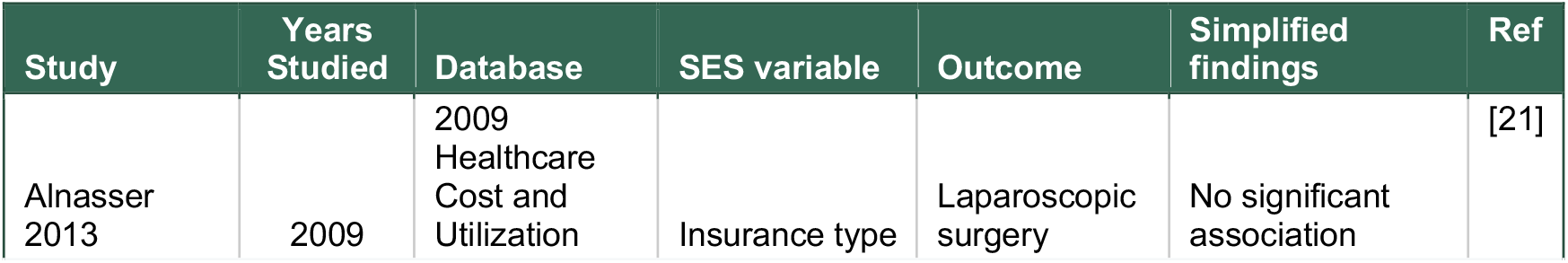
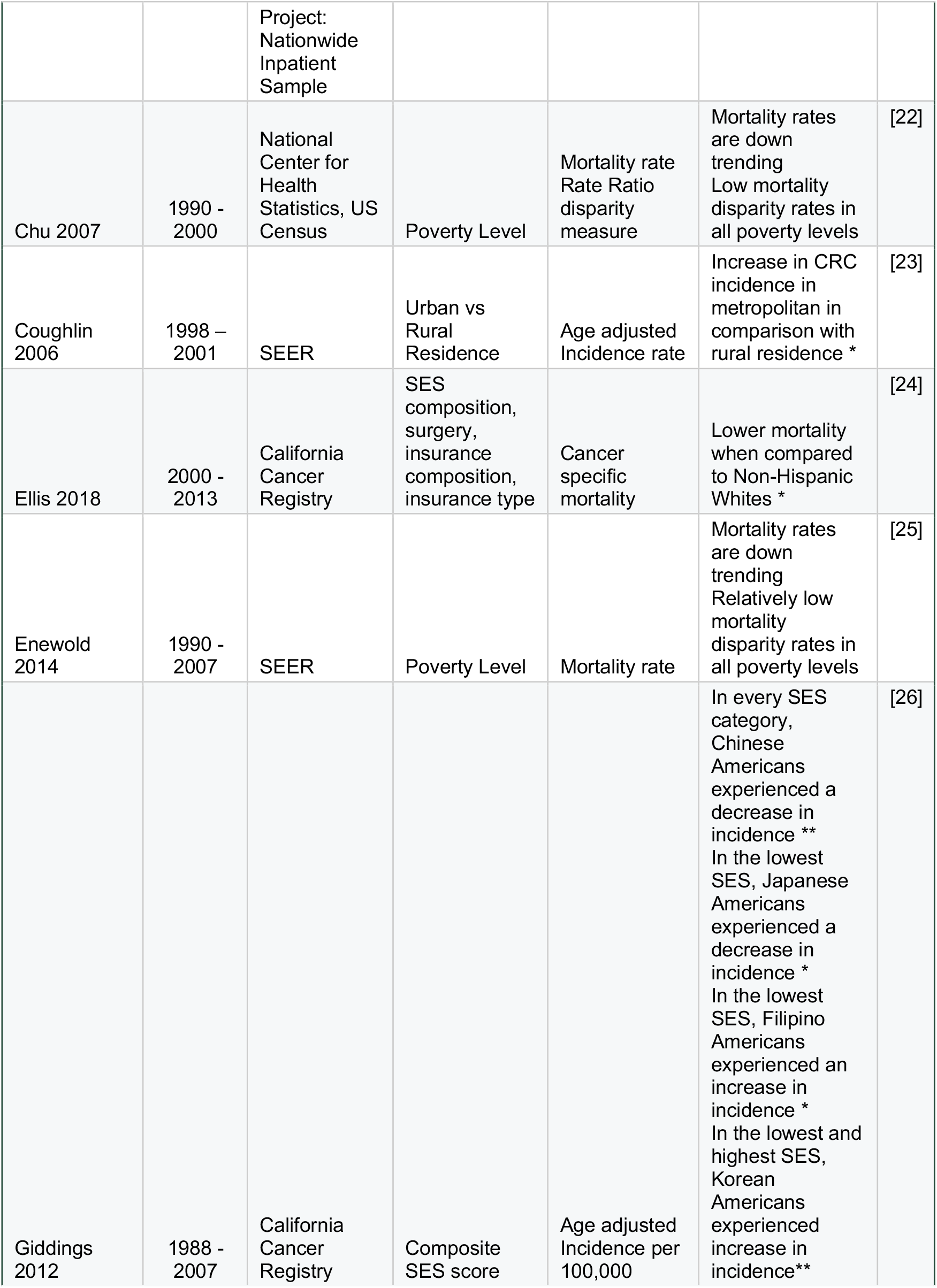
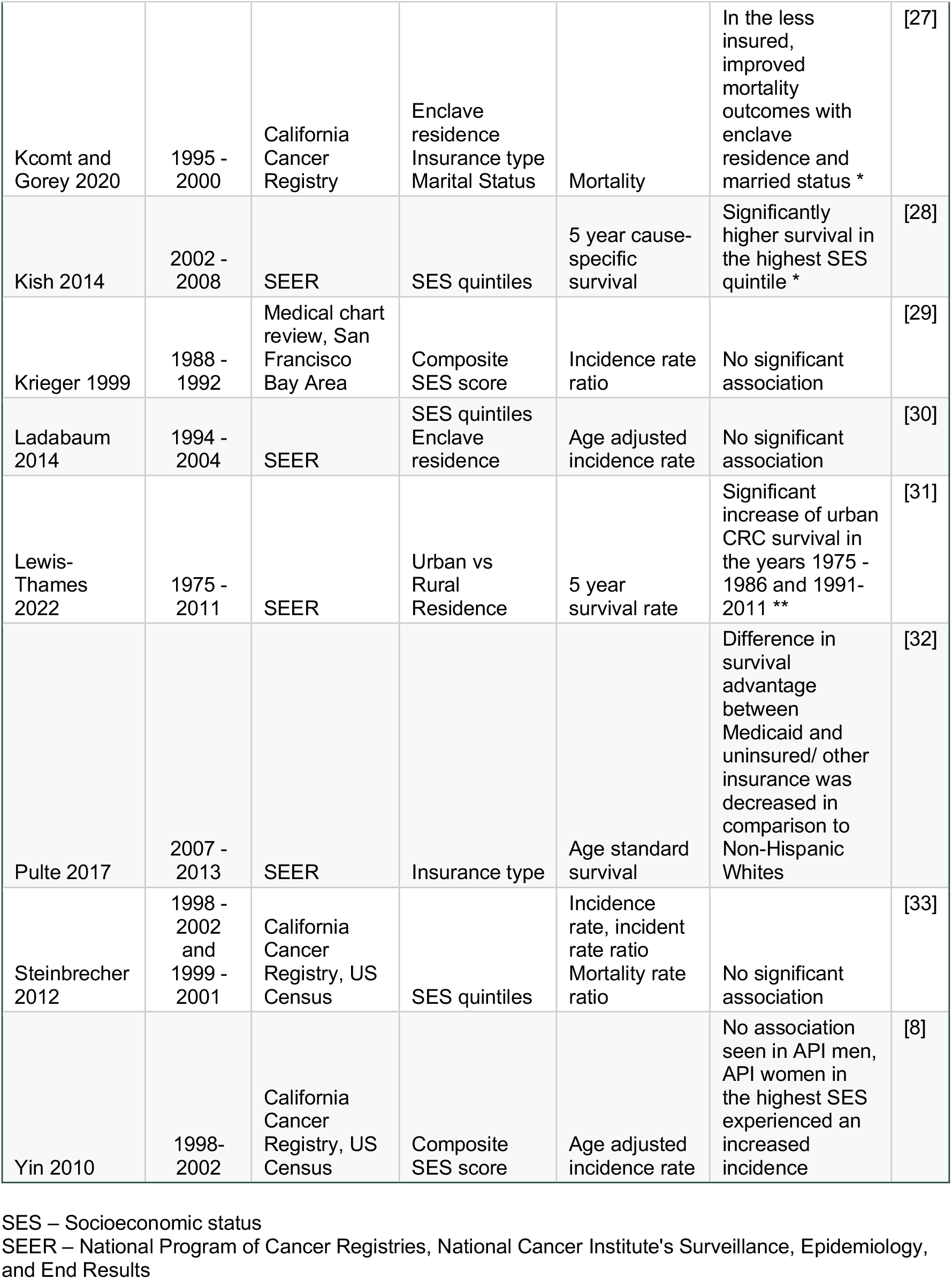
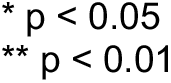

### Incidence

Six studies measured incidence as an outcome. One study found a significant increase in incidence in urban areas compared to rural in the API population [23]. When investigating incidence trends among Asian American subgroups, Giddings et. al found that Chinese Americans experienced a significant decrease in every SES category; Japanese Americans experienced a significant decrease in the lowest SES; and Korean Americans and Filipino Americans experienced a significant increase in incidence both in the lowest and in the highest SES categories [26]. The other 4 studies concluded that there was no association or variable response to SES measurements [22,24,25,33,34].

### Mortality and Survival

Four out of the five studies stated that mortality rates are low in the API group, despite changes in SES factors [22,24,25,33]. One study revealed that marital status and the presence of an ethnic enclave improved mortality, and Chinese Americans benefited more from enclaves than other ethnicities [27]. Similarly, studies found an increased survival rate in the API group regardless of changes in SES [28,31,32].

## Discussion

This is the first systematic review to assess the impact of socioeconomic factors on CRC outcomes, specifically within the Asian American population. Existing literature predominantly reports that the incidence and mortality rates associated with CRC are either low or experiencing a downward trend across various SES strata among Asian Pacific Islanders (API). Our findings align with the prevailing epidemiological consensus indicating that Asian Americans exhibit improved CRC outcomes relative to other racial and ethnic groups. Nonetheless, this review also revealed significant heterogeneity in CRC outcomes when analyzing the population as a consolidated entity, as illustrated in Figure 2. Notably, there is emerging evidence suggesting that the incidence of CRC is on the rise in certain subgroups of Asian Americans, contrasting the national trends [30,34].

**Figure 2.**
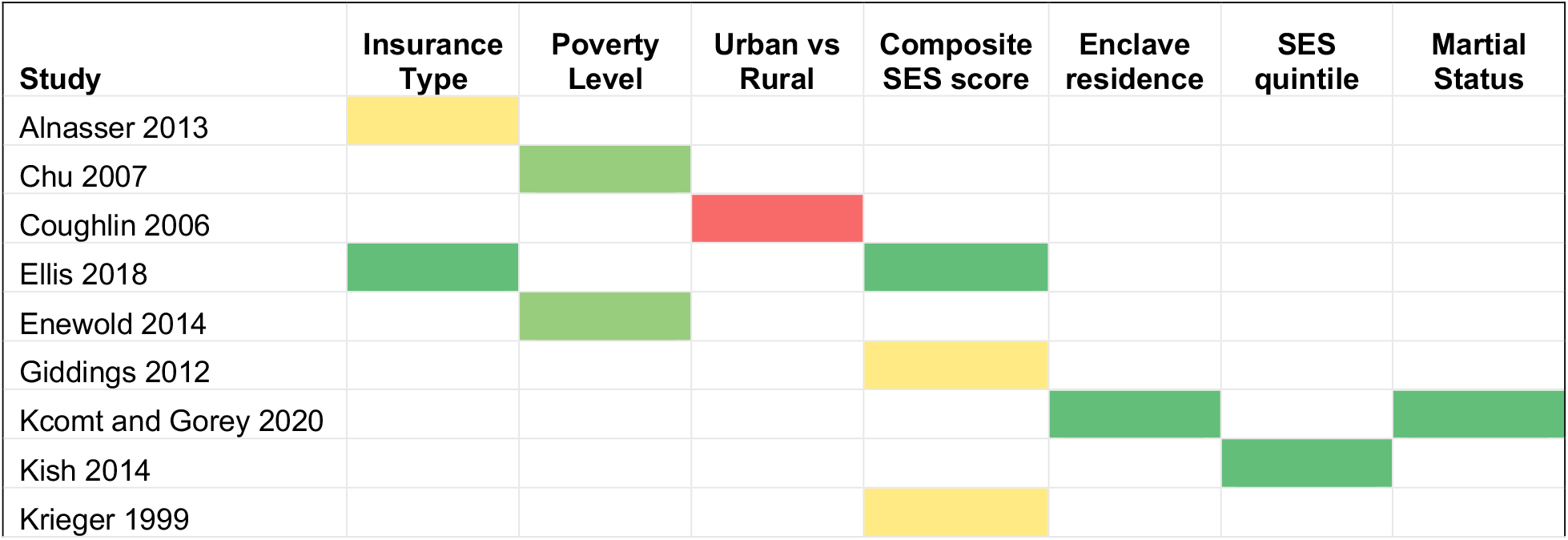
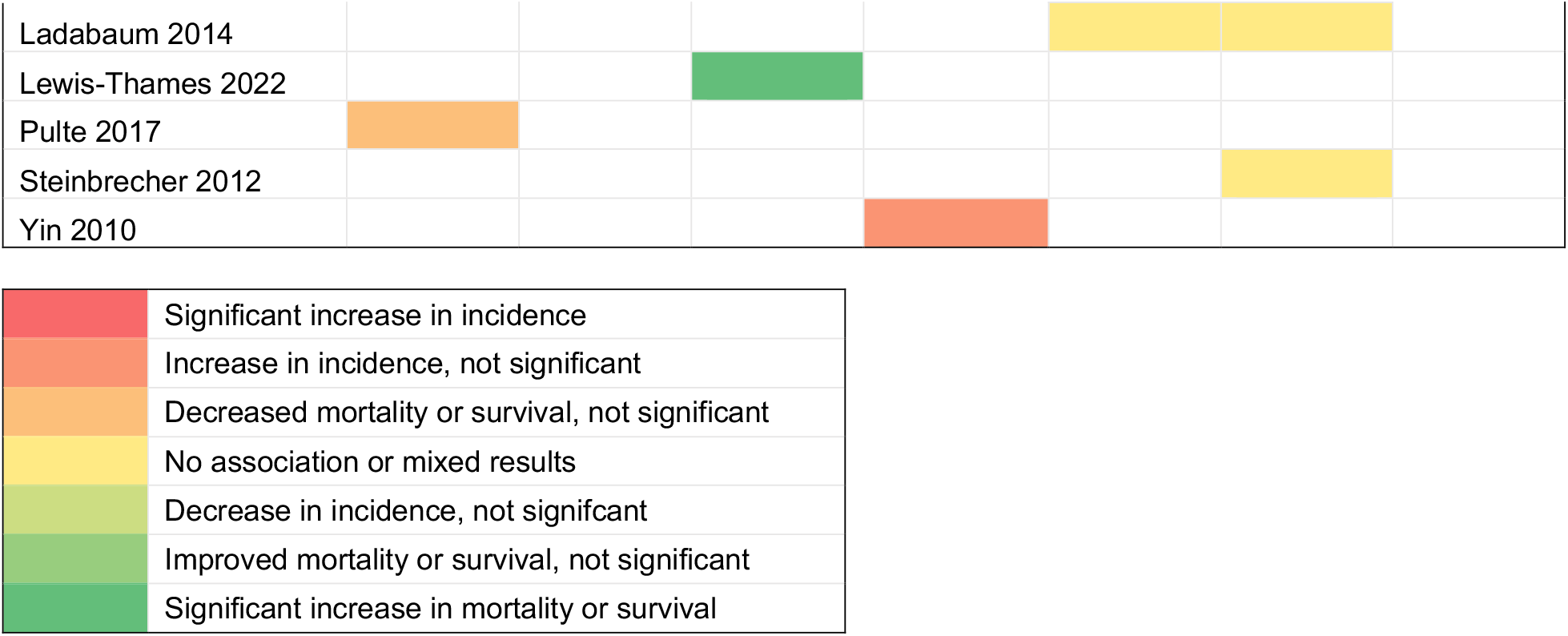
Heat Map reflecting our study results.

This review underscores that the designation of a singular monolithic “Asian” category is misleading in epidemiological assessments of CRC. When analyzed as distinct ethnic or racial identities, there is notable heterogeneity in incidence trends and outcomes within Asian populations [15]. Specifically, while CRC prevalence has exhibited an upward trajectory among Asian Americans, national trends indicate a general decline [34]. A 1998 report highlighted that among Asian American subgroups, the highest incidence rates were observed in Japanese men (64.1 per 100,000) and Alaska Natives (79.7 per 100,000) [35]. Furthermore, a 2013 investigation illuminated increasing incidence rates among Korean and Native Hawaiian populations [36]. Notably, a population-based study revealed that Laotian, Samoan, and Vietnamese men were significantly less likely to receive diagnoses at earlier stages of CRC [37]. This disparity emphasizes the necessity for disaggregated data to depict the epidemiology of CRC across diverse Asian subpopulations accurately.

Secondly, mortality rates from CRC also varied among ethnic subgroups, with Native Hawaiians and Southeast Asians having the greatest risk of mortality from CRC, but Chinese, Japanese, and Indians/Pakistanis had a lower risk [16,17]. Japanese Americans in Hawaii also were reported to have improved survival rates in the long term [38]. Additionally, foreign-born Asian Americans are more likely to have increased mortality from CRC [39]. However, Chinese men and Filipino persons had lower CRC incidence while foreign-born Japanese had a higher incidence [30,40]. These studies highlight the heterogeneity in CRC statistics among Asian American groups. Some groups do demonstrate more favorable survival outcomes, but other groups are more at risk-a statistic that is overseen if all Asian Americans were treated monolithically [16]. Additionally, despite national improvement in screening that has contributed to an overall decrease in mortality, screening rates among Asian American groups have been inadequate [41,42].

Finally, the widening of SES disparity in the last several decades has correlated with mortality from all cancers and extends to cardiovascular disease as well [18]. For CRC specifically, it has been demonstrated that low SES, including poverty, lack of education, lack of social support, and social isolation are associated with poorer survival in CRC [18,43]. Low SES also correlates to a lack of health insurance and access to care, which is a barrier to receiving appropriate screening, leading to later-stage disease at diagnosis and higher mortality [18]. However, SES and its relationship to disparities among race and ethnicity is complex, and studies have shown a variable association with outcomes [28,29]. Further, studies suggest that the extent to which SES factors, such as private health insurance or marital status, improve cancer survival can differ among race/ethnicity [11,44]. It is essential to have a more detailed ethnic classification of the API population as well as to learn about the genetic background of each ethnicity, which could lead to a more precise assessment and potential treatment of CRC based on molecular mechanisms [45,46].

Importantly, despite the high incidence of CRC in Asian Americans, this group has a lower rate of CRC screening compared to Whites and African Americans. However, it has been reported that only 50% of Asian individuals have up-to-date colonoscopies compared to 61% of White and African Americans [1.41.47]. Several cultural factors contribute to this disparity, including a lack of CRC knowledge, less acculturation, cancer-related fatalism, poor English language proficiency, embarrassment about screenings, and limited social support, to name a few [48-52]. To ensure that Asian Americans receive the appropriate CRC screenings, it is important to have culturally and socially appropriate counseling and guidance. This population has diverse cultural beliefs and practices and thus requires individualized support to overcome the barriers to screening [49.51].

The limitations of this review are as follows: (1) We conducted a literature search for potential studies on the databases used (Method): (2) The publication bias of the articles is considered low in studies measuring prevalence, incidence, and mortality studies, and thus we did not assess the bias. (3) Studies that focused on specific Asian ethnic groups were limited. We suggest that more studies considering heterogeneous groups are carried out as the Asian American population continues to grow and diversify.

## Concluding remarks

The review collectively illustrates a concerning increase in CRC incidence among various Asian American ethnic groups. These disparities are likely attributable to a combination of heterogeneous factors, including inadequate screening rates, insufficient educational outreach regarding CRC, and prevalent cultural barriers. It is essential to pursue further research to elucidate these underlying mechanisms. Consequently, this review advocates for a more detailed categorization of the API ethnic populations, moving beyond the monolithic classification of “Asian.” Furthermore, it emphasizes the urgent need for preventative CRC screening initiatives within API communities, which currently exhibit lower screening rates compared to other demographic groups.

## Data Availability

All data produced in the present work are contained in the manuscript.

## Abbreviations

(API): Asian American and Pacific Islander
(CRC): Colorectal cancer
(SES): Socioeconomic status

## Acknowledgments

We would like to express our gratitude to the members of the Asian Pacific American Medical Student Association (APAMSA) for their valuable contributions to our discussion. We also extend our thanks to the members of the Murakami lab who helped proofread this review. Additionally, we utilized grammar-check software that incorporates AI technology (https://app.grammarly.com; accessed on 7 January 2025) for English editing purposes only.

## References

1. Siegel RL, Wagle NS, Cercek A, Smith RA, Jemal A. Colorectal cancer statistics, 2023. CA Cancer J Clin. May-Jun 2023;73(3):233–254. doi:10.3322/caac.21772

2. Miller KD, Nogueira L, Devasia T, et al. Cancer treatment and survivorship statistics, 2022. CA Cancer J Clin. Sep 2022;72(5):409–436. doi:10.3322/caac.21731

3. Islami F, Goding Sauer A, Miller KD, et al. Proportion and number of cancer cases and deaths attributable to potentially modifiable risk factors in the United States. CA Cancer J Clin. Jan 2018;68(1):31–54. doi:10.3322/caac.21440

4. Lin JS, Perdue LA, Henrikson NB, Bean SI, Blasi PR. Screening for Colorectal Cancer: Updated Evidence Report and Systematic Review for the US Preventive Services Task Force. Jama. May 18 2021;325(19):1978–1998. doi:10.1001/jama.2021.4417

5. Gu M, Thapa S. Colorectal cancer in the United States and a review of its heterogeneity among Asian American subgroups. Asia Pac J Clin Oncol. Aug 2020;16(4):193–200. doi:10.1111/ajco.13324

6. Monte L, Shin H. 20.6 Million People in the U.S. Identify as Asian, Native Hawaiian or Pacific Islander. August 3, 2023. Accessed August 3, 2023. https://www.census.gov/library/stories/2022/05/aanhpi-population-diverse-geographically-dispersed.html#:~:text=originated%20in%20Asia.-,Race,in%20combination%20with%20another%20race.

7. Budiman A, Ruiz N. Asian Americans are the fastest-growing racial or ethnic group in the U.S. Internet. August 3, 2023. Accessed August 3, 2023. https://www.pewresearch.org/short-reads/2021/04/09/asian-americans-are-the-fastest-growing-racial-or-ethnic-group-in-the-u-s/

8. Yin D, Morris C, Allen M, Cress R, Bates J, Liu L. Does socioeconomic disparity in cancer incidence vary across racial/ethnic groups? Cancer Causes Control. Oct 2010;21(10):1721–30. doi:10.1007/s10552-010-9601-y

9. Lee RJ, Madan RA, Kim J, Posadas EM, Yu EY. Disparities in Cancer Care and the Asian American Population. Oncologist. Jun 2021;26(6):453–460. doi:10.1002/onco.13748

10. Mulhern KC, Wahl TS, Goss LE, et al. Reduced disparities and improved surgical outcomes for Asian Americans with colorectal cancer. J Surg Res. Oct 2017;218:23–28. doi:10.1016/j.jss.2017.05.036

11. Pan HY, Walker GV, Grant SR, et al. Insurance Status and Racial Disparities in Cancer-Specific Mortality in the United States: A Population-Based Analysis. Cancer Epidemiol Biomarkers Prev. Jun 2017;26(6):869–875. doi:10.1158/1055-9965.Epi-16-0976

12. Hashiguchi Y, Hase K, Ueno H, et al. Impact of race/ethnicity on prognosis in patients who underwent surgery for colon cancer: analysis for white, African, and East Asian Americans. Ann Surg Oncol. May 2012;19(5):1517–28. doi:10.1245/s10434-011-2113-5

13. Alshareef SH, Alsobaie NA, Aldeheshi SA, Alturki ST, Zevallos JC, Barengo NC. Association between Race and Cancer-Related Mortality among Patients with Colorectal Cancer in the United States: A Retrospective Cohort Study. Int J Environ Res Public Health. Jan 16 2019;16(2)doi:10.3390/ijerph16020240

14. Oh DL, Santiago-Rodríguez EJ, Canchola AJ, Ellis L, Tao L, Gomez SL. Changes in Colorectal Cancer 5-Year Survival Disparities in California, 1997-2014. Cancer Epidemiol Biomarkers Prev. Jun 2020;29(6):1154–1161. doi:10.1158/1055-9965.Epi-19-1544

15. Thompson CA, Gomez SL, Hastings KG, et al. The Burden of Cancer in Asian Americans: A Report of National Mortality Trends by Asian Ethnicity. Cancer Epidemiol Biomarkers Prev. Oct 2016;25(10):1371–1382. doi:10.1158/1055-9965.Epi-16-0167

16. Medina HN, Callahan KE, Morris CR, Thompson CA, Siweya A, Pinheiro PS. Cancer Mortality Disparities among Asian American and Native Hawaiian/Pacific Islander Populations in California. Cancer Epidemiol Biomarkers Prev. Jul 2021;30(7):1387–1396. doi:10.1158/1055-9965.Epi-20-1528

17. Chien C, Morimoto LM, Tom J, Li CI. Differences in colorectal carcinoma stage and survival by race and ethnicity. Cancer. Aug 1 2005;104(3):629–39. doi:10.1002/cncr.21204

18. Singh GK, Jemal A. Socioeconomic and Racial/Ethnic Disparities in Cancer Mortality, Incidence, and Survival in the United States, 1950–2014: Over Six Decades of Changing Patterns and Widening Inequalities. Journal of Environmental and Public Health. 2017/03/20 2017;2017:2819372. doi:10.1155/2017/2819372

19. Page MJ, McKenzie JE, Bossuyt PM, et al. The PRISMA 2020 statement: an updated guideline for reporting systematic reviews. Bmj. Mar 29 2021;372:n71. doi:10.1136/bmj.n71

20. Covidence systematic review software. 2023. Accessed 2024. https://www.covidence.org.

21. Alnasser M, Schneider EB, Gearhart SL, et al. National disparities in laparoscopic colorectal procedures for colon cancer. Surg Endosc. Jan 2014;28(1):49–57. doi:10.1007/s00464-013-3160-8

22. Chu KC, Miller BA, Springfield SA. Measures of racial/ethnic health disparities in cancer mortality rates and the influence of socioeconomic status. J Natl Med Assoc. Oct 2007;99(10):1092–100, 1102-4.

23. Coughlin SS, Richards TB, Thompson T, et al. Rural/nonrural differences in colorectal cancer incidence in the United States, 1998-2001. Cancer. Sep 1 2006;107(5 Suppl):1181–8. doi:10.1002/cncr.22015

24. Ellis L, Canchola AJ, Spiegel D, Ladabaum U, Haile R, Gomez SL. Racial and Ethnic Disparities in Cancer Survival: The Contribution of Tumor, Sociodemographic, Institutional, and Neighborhood Characteristics. J Clin Oncol. Jan 1 2018;36(1):25–33. doi:10.1200/jco.2017.74.2049

25. Enewold L, Horner MJ, Shriver CD, Zhu K. Socioeconomic disparities in colorectal cancer mortality in the United States, 1990-2007. J Community Health. Aug 2014;39(4):760–6. doi:10.1007/s10900-014-9824-z

26. Giddings BH, Kwong SL, Parikh-Patel A, Bates JH, Snipes KP. Going against the tide: increasing incidence of colorectal cancer among Koreans, Filipinos, and South Asians in California, 1988-2007. Cancer Causes Control. May 2012;23(5):691–702. doi:10.1007/s10552-012-9937-6

27. Kcomt L, Gorey KM. Chinese enclave protections among married Chinese American women: exploratory secondary analysis of colon cancer survival. Ethn Health. Nov 2020;25(8):1089–1102. doi:10.1080/13557858.2018.1493432

28. Kish JK, Yu M, Percy-Laurry A, Altekruse SF. Racial and ethnic disparities in cancer survival by neighborhood socioeconomic status in Surveillance, Epidemiology, and End Results (SEER) Registries. J Natl Cancer Inst Monogr. Nov 2014;2014(49):236–43. doi:10.1093/jncimonographs/lgu020

29. Krieger N, Quesenberry C, Jr., Peng T, et al. Social class, race/ethnicity, and incidence of breast, cervix, colon, lung, and prostate cancer among Asian, Black, Hispanic, and White residents of the San Francisco Bay Area, 1988-92 (United States). Cancer Causes Control. Dec 1999;10(6):525–37. doi:10.1023/a:1008950210967

30. Ladabaum U, Clarke CA, Press DJ, et al. Colorectal cancer incidence in Asian populations in California: effect of nativity and neighborhood-level factors. Am J Gastroenterol. Apr 2014;109(4):579–88. doi:10.1038/ajg.2013.488

31. Lewis-Thames MW, Langston ME, Khan S, et al. Racial and Ethnic Differences in Rural-Urban Trends in 5-Year Survival of Patients With Lung, Prostate, Breast, and Colorectal Cancers: 1975-2011 Surveillance, Epidemiology, and End Results (SEER). JAMA Netw Open. May 2 2022;5(5):e2212246. doi:10.1001/jamanetworkopen.2022.12246

32. Pulte D, Jansen L, Brenner H. Social disparities in survival after diagnosis with colorectal cancer: Contribution of race and insurance status. Cancer Epidemiol. Jun 2017;48:41–47. doi:10.1016/j.canep.2017.03.004

33. Steinbrecher A, Fish K, Clarke CA, West DW, Gomez SL, Cheng I. Examining the association between socioeconomic status and invasive colorectal cancer incidence and mortality in California. Cancer Epidemiol Biomarkers Prev. Oct 2012;21(10):1814–22. doi:10.1158/1055-9965.Epi-12-0659

34. Jandova J, Ohlson E, Torres BSM, et al. Racial disparities and socioeconomic status in the incidence of colorectal cancer in Arizona. Am J Surg. Sep 2016;212(3):485–92. doi:10.1016/j.amjsurg.2015.08.024

35. Parker SL, Davis KJ, Wingo PA, Ries LA, Heath CW, Jr. Cancer statistics by race and ethnicity. CA Cancer J Clin. Jan-Feb 1998;48(1):31–48. doi:10.3322/canjclin.48.1.31

36. Gomez SL, Noone AM, Lichtensztajn DY, et al. Cancer incidence trends among Asian American populations in the United States, 1990-2008. J Natl Cancer Inst. Aug 7 2013;105(15):1096–110. doi:10.1093/jnci/djt157

37. Miller BA, Chu KC, Hankey BF, Ries LA. Cancer incidence and mortality patterns among specific Asian and Pacific Islander populations in the U.S. Cancer Causes Control. Apr 2008;19(3):227–56. doi:10.1007/s10552-007-9088-3

38. Hata M, Sakamoto K, Doneza J, et al. Improvement of long-term survival of colorectal cancer in Japanese-Americans of Hawaii from 1990 to 2001. Int J Clin Oncol. Dec 2010;15(6):559–64. doi:10.1007/s10147-010-0103-4

39. Choe JH, Koepsell TD, Heagerty PJ, Taylor VM. Colorectal cancer among Asians and Pacific Islanders in the U.S.: survival disadvantage for the foreign-born. Cancer Detect Prev. 2005;29(4):361–8. doi:10.1016/j.cdp.2005.06.002

40. Redaniel MT, Laudico A, Mirasol-Lumague MR, et al. Cancer survival discrepancies in developed and developing countries: comparisons between the Philippines and the United States. Br J Cancer. Mar 10 2009;100(5):858–62. doi:10.1038/sj.bjc.6604945

41. Domingo JB, Chen JJ, Braun KL. Colorectal Cancer Screening Compliance among Asian and Pacific Islander Americans. J Immigr Minor Health. Jun 2018;20(3):584–593. doi:10.1007/s10903-017-0576-6

42. Zauber AG, Winawer SJ, O’Brien MJ, et al. Colonoscopic polypectomy and long-term prevention of colorectal-cancer deaths. N Engl J Med. Feb 23 2012;366(8):687–96. doi:10.1056/NEJMoa1100370

43. Coughlin SS. Social determinants of colorectal cancer risk, stage, and survival: a systematic review. Int J Colorectal Dis. Jun 2020;35(6):985–995. doi:10.1007/s00384-020-03585-z

44. Aizer AA, Chen MH, McCarthy EP, et al. Marital status and survival in patients with cancer. J Clin Oncol. Nov 1 2013;31(31):3869–76. doi:10.1200/jco.2013.49.6489

45. Kim J, Kang SJ, Jo N, Kim SJ, Jang S. Cancer prognosis using base excision repair genes. Mol Cells. 2025 Jan 17;48(2):100186. doi: 10.1016/j.mocell.2025.100186.

46. Badial K, Lacayo P, Murakami S. Biology of Healthy Aging: Biological Hallmarks of Stress Resistance Related and Unrelated to Longevity in Humans. Int J Mol Sci. 2024 Sep 29;25(19):10493. doi: 10.3390/ijms251910493.

47. Hwang H. Colorectal cancer screening among Asian Americans. Asian Pac J Cancer Prev. 2013;14(7):4025–32. doi:10.7314/apjcp.2013.14.7.4025

48. Jin SW, Yoon YJ. Barriers and facilitators to colorectal cancer screening among older Korean Americans: A focus group study. Soc Work Health Care. Oct-Dec 2020;59(9-10):668–680. doi:10.1080/00981389.2020.1852359

49. Jun J, Nan X. Determinants of Cancer Screening Disparities Among Asian Americans: A Systematic Review of Public Health Surveys. J Cancer Educ. Aug 2018;33(4):757–768. doi:10.1007/s13187-017-1211-x

50. Juon HS, Guo J, Kim J, Lee S. Predictors of Colorectal Cancer Knowledge and Screening Among Asian Americans Aged 50-75 years old. J Racial Ethn Health Disparities. Jun 2018;5(3):545–552. doi:10.1007/s40615-017-0398-1

51. Kim SB. Unraveling the Determinants to Colorectal Cancer Screening Among Asian Americans: a Systematic Literature Review. J Racial Ethn Health Disparities. Aug 2018;5(4):683–699. doi:10.1007/s40615-017-0413-6

52. Oh KM, Jacobsen KH. Colorectal cancer screening among Korean Americans: a systematic review. J Community Health. Apr 2014;39(2):193–200. doi:10.1007/s10900-013-9758-x

